# Profile of Patients with Sleep Complaints in a Secondary Care Health Institution in the City of Campinas, São Paulo State

**DOI:** 10.1101/2023.04.07.23286749

**Authors:** Paulo Afonso Mei

**Affiliations:** São Leopoldo Mandic Medical College, Campinas-SP, Brazil

**Keywords:** Sleep Medicine, Anxiety, Depression, Quality of Life, Sleep Initiation and Maintenance Disorders, Sleepiness

## Abstract

**Objective:** We report data of the profile of patients who sought our outpatient clinic for sleep disorders of the Faculty of Medicine of São Leopoldo Mandic Medical School, during the Week of Sleep, that took place in March, 2022.

**Methods:** All participants responded to a form designed by the researcher, as well as to commonly used questionnaires in Sleep Research, such as the Epworth Sleepiness Scale (ESS), the Insomnia Severity Index (ISI), the Hospital Anxiety and Depression Scale, the STOP-BANG questionnaire for screening of apnea, and the WHO Quality of Life abbreviated questionnaire (WHOQOL-BREF).

**Results:** 30 patients were evaluated, mean age 51.8 (± 14), 63% women. Main complaints were insomnia (63%), snore/apnea (23%) and excessive daytime sleepiness (10%). ESS scores were significantly higher among men, daytime nappers and alarm clock users. ISI scores were importantly associated with those who woke 2 or more times per night, while physically active, non-obese, normotensive people and those with Modified Mallampatti (MM) classes I and II scored significantly less in STOP-BANG. Obese, MM classes III-IV and people with abnormal cervical and abdominal circumferences performed markedly worse in Domain 1, while women, older people and alarm clock users went worse in Domain 4 of WHOQOL-BREF.

**Discussion:** One or more sleep disturbances were detected in all seekers of Sleep Care, emphasizing the importance of these types of action.

## INTRODUCTION

Concerns with the sleep health are a matter of growing relevance within the medical community, especially regarding guidelines for the management of sleep issues as a part of not only public health, but also economy^1^, as workers with bad sleep tend to be more absent, which implies shortfalls in supply from the workforce of enterprises. Also, the actual post-pandemic scenario has shown that the rise of sleep issues derived directly or indirectly from the SARS-CoV 2 virus (causer of COVID-19) has not yet returned to pre-pandemic levels^2^.

In this sense, this paper describes the profile of patients that sought the outpatient clinic for sleep disorders of our faculty, during a campaign promoted by our university on March 2022, entitled “*Semana do Sono 2022*” [Sleep Week 2022], as well as the more remarking associations with common sleep questionnaire scores we obtained.

## MATERIAL AND METHODS

We provided 50 appointment times for medical consultation, free of charge, at the Sleep Medicine Disorders outpatient clinic, located inside the campus of São Leopoldo Mandic Medical School, in the city of Campinas, São Paulo State, Brazil, which serves a metropolitan region of roughly 2 million inhabitants and functions in a mixed model of providing both private and public health consultations. Prior to the availability of care, there was broad advertisement of the campaign by a press release from our journalism department^3^, which was replicated by a reasonable amount of local medias, mainly FM radios and internet portals.^4 5 6^

Criteria for inclusion were to be 10 years of age or older, with a self or caregiver complaint of bad sleep, regardless of previous treatments on the matter. There were no restrictions other than age, as we intended to faithfully portray by sample the profile of people with sleep issues in our region.

Before recruitment, ethical approval from the board of directors of the clinic was obtained. Patients were required to schedule appointments through phone call or web message. Patients younger than 18 years old or in face of limitation of comprehension were required to be accompanied by a caregiver or progenitor.

At arrival, patients or tutors were required to read and sign an informed consent, and fill out the following questionnaires: Epworth Sleepiness Scale^7^ (ESS), Insomnia Severity Index^8^ (ISI), Hospital Anxiety and Depression Scale^9^ (HADS), STOP-BANG^10^ questionnaire for triage of possible apnoea sufferers, and Abbreviated World Health Organization Quality of Life survey^11^ (WHOQOL-BREF), which is divided into four domains (Ds) – D1 (Physical wellbeing), D2 (Psychological wellbeing), D3 (Social wellbeing) and D4 (Environmental wellbeing). All questionnaires were presented in the corresponding, validated Portuguese versions^12 13 14 15 16^.

After filling out the forms, patients underwent consultation by the head of the clinic (the author) or by medical students and interns, always under direct guidance by the former.

On anamnesis, we sought to identify conditions and behaviours that were known to be related to sleep issues, such as type of shift, presence of comorbidities, time went to bed, time that fell asleep, average of times awaken at night, time of last wake-up, time that arose from bed, use of alarm clock or smartwatches on a daily basis, consume of caffeine, alcohol and tobacco, if participants exercised - frequency and time of day, medications in current use and previously used, if they were infected by the SARS-CoV-2 coronavirus (COVID-19) and time of infection, if they ever did sleep study by polysomnography (PSG), etc.

We performed a general physical exam in all patients, as at the time of the research students were facing normal rotations and always were required to do so, but we emphasized attributes that could interfere with sleep, such as height and weight, to further calculate the body mass index (BMI), Blood Pressure, phenotype of dental occlusion (retrognathic, normognathic or prognathic), size of tonsils (Grades 0 to IV), the Modified Mallampati Score^17^ (MM) - which we further divided in patients with favourable airway, if a MM Class I or II was present, or unfavourable airway, in cases of MM Classes III and IV. We also measured neck (NC) and waist (WC) circumferences.

## RESULTS

From the 50 appointments, 30 patients attended in the schedule time (40% of no-show). Mean age was 58.1 years old (SD ± 14). As the youngest patient was 23, there were no paediatric subjects.

The demographic profile of our sample is portrayed in **Table 1**, where it is evident that the vast majority (87%) comprised insomnia or snore/witnessed apnoea as the main complaints.

**Table 1.** Characteristics of the sample studied. *N = 21

| Demography | N | % |
| --- | --- | --- |
| Total (Attendees) | 30 | 100 |
| Mean Age (± SD) | 51.8 ± 14 | 13.9 |
| • Minimal Age | 23 | — |
| • Maximal Age | 78 | — |
| • Mean | 54 |  |
| • Q1 | 40.3 |  |
| • Q2 | 53.9 |  |
| • Q3 | 61.1 |  |
| • Q4 | 78.0 |  |
| Women | 19 | 63.3 |
| Men | 11 | 36.7 |
| Type of Work Shift | N | % |
| Exclusively diurnal (between 8:00 a.m. and 06:00 p.m.) | 26 | 86.7 |
| Extended Diurnal (till evening) | 2 | 6.7 |
| Mixed Nocturnal/Diurnal | 2 | 6.7 |
| Exclusively Nocturnal | 0 | 0 |
| Main complaint/Axis | N | % |
| Insomnia | 19 | 63.3 |
| Snore/Apnoea | 7 | 23.3 |
| Daytime Sleepiness | 3 | 10.0 |
| Rhythm Disorders | 0 | 0 |
| Movement Disorders | 0 | 0 |
| Parasomnias | 0 | 0 |
| Other | 1 | 3.3 |
| Comorbidities (Self referred) and Habits | N | % |
| Hypertension | 16 | 53.3 |
| Dyslipidaemia | 5 | 16.7 |
| Had COVID-19 | 8 | 26.7 |
| Depression | 8 | 26.7 |
| Anxiety | 22 | 73.3 |
| DM or Pre-DM | 6 | 20.0 |
| Smoking | 3 | 10.0 |
| Alcohol drinking | 17 | 56.7 |
| Exercise | 13 | 43.3 |
| • Only diurnal | 8 | 26.7 |
| • Only nocturnal | 2 | 6.7 |
| • Diurnal and nocturnal | 3 | 10.0 |
| • Only aerobic | 8 | 26.7 |
| • Only anaerobic | 0 | 0 |
| • Aerobic + Anaerobic | 5 | 16.7 |

**Table 1 - Characteristics of the sample studied**
| Use of Smartwatch | 5 | 16.7 |
| --- | --- | --- |
| Use of Alarm Clock | 12 | 40.0 |
| Napping | 7* | 33.3* |
| • Morning napping | 6 |  |
| • napping ≤ 20 minutes | 1 | |
| • napping $>$ 20 minutes | 5 | |
| • Afternoon napping | 2 |  |
| • napping $>$ 20 min | 1 | |
| • napping $>$ 20 min | 1 | |
| Physical Exam Data | N | SD |
| Mean BMI | 30.8 | 7.4 |
| Mean neck circumference | 38.9 | 4.4 |
| Mean abdominal circumference | 105.3 | 17.2 |
| Mean Systolic Pressure | 131.7 | 19.2 |
| Mean Diastolic Pressure | 84.0 | 12.5 |
| Previous Investigations |  |  |
| Ever had Polysomnography (PSG) made | 4 | 13.3 |
| • PSG made in the last 5 years | 2 | 6.7 |

Only one male patient’s main complaint did not fall in a category that could correspond to a sleep disorder diagnosis, according to the International Classification of Sleep Disorders, third edition^18^ (ICSD-3), as he described paroxysmal, nocturnal events that, by our judgment, filled criteria for panic disorder, according to the 11^th^ International Classification of Diseases^19^ (ICD-11 code 6B01), but nonetheless was included in the analysis, as the events had negative consequences both on sleep and daytime functioning.

**Tables 2 and 3** present data from our statistical analyses. **Table 2** comprises the mean scores of the aforementioned questionnaires for the whole sample and by groups. In **Table 3** we explored analyses of categorical attributes regarding excessive daytime sleepiness (defined by ESS ≥ 9 points) and probable insomniacs (ISI ≥ 15 points). The best cut-off scores of both scales remains disputed ^20 21^, being our choices of values supported by current literature ^22^.

**Table 2.** Mean of tested scores according to different groups or conditions and independent t-test results for comparison of categorical vs. continuous variables –. * - N = 21, ** N = 28, ^d^ – t-test for independent samples; ‡ - waist circumference considered abnormal when ≥ 91 cm in women and ≥ 98 cm in men; ◊ - neck circumference considered abnormal when ≥ 38 cm in women and ≥ 41 cm in men; D1, D2, D3 e D4 — Domains 1 (Physical), 2 (Psycho), 3 (Social) and 4 (Environmental) of WHOQOL-BREF - World Health Organization Quality of Life - Brief version, ESS - Epworth Sleepiness Scale, HADS-A e HADS-D — Hospital Anxiety (A) and Depression (D) Scale, SH — Sleep Hygiene, ISI — Insomnia Severity Index, MM — Modified Mallampati Score

|  |  |  |  |  |  |  |  |  |  |  | WHOQOL-BREF |  |  |  |  |  |  |  |
| --- | --- | --- | --- | --- | --- | --- | --- | --- | --- | --- | --- | --- | --- | --- | --- | --- | --- | --- |
| Condition/Group | ESS |  | HADS-A |  | HADS-D |  | ISI |  | STOP-BANG |  | D1 |  | D2 |  | D3 |  | D4 |  |
|  | Mean | p | Mean | p | Mean | p | Mean | p | Mean | p | Mean | p | Mean | p | Mean | p | Mean | p |
| Women | 6.47 | .013 | 11.26 | .106 | 8.74 | .236 | 17.24 | .157 | 3.18 | .070 | 48.6 | .473 | 53.8 | .246 | 67.8 | .581 | 55.4 | .045 |
| Men | 11.55 |  | 8.82 |  | 6.91 |  | 14.36 |  | 4.55 |  | 53.0 |  | 60.0 |  | 65.9 |  | 64.9 |  |
| Younger half | 8.46 | .998 | 10.43 | .938 | 8.43 | .652 | 16.38 | .798 | 3.08 | .110 | 52.4 | .498 | 58.1 | .466 | 66.6 | .373 | 64.2 | .027 |
| Older half | 8.47 |  | 10.31 |  | 7.75 |  | 15.87 |  | 4.27 |  | 48.3 |  | 54.3 |  | 60.1 |  | 54.1 |  |
| Had COVID before | 8.14 | .861 | 11.25 | .473 | 9.63 | .205 | 17.57 | .399 | 3.57 | .829 | 48.6 | .463 | 55.6 | .840 | 62.2 | .807 | 55.4 | .443 |
| Never had COVID | 8.57 |  | 10.05 |  | 7.50 |  | 15.62 |  | 3.76 |  | 53.4 |  | 56.9 |  | 64.3 |  | 59.9 |  |
| Exercises | 7.30 | .409 | 11.17 | .378 | 8.83 | .403 | 15.60 | .708 | 2.90 | .049<sup>d</sup> | 54.7 | .274 | 52.6 | .306 | 62.5 | .856 | 57.0 | .505 |
| Sedentary | 9.11 |  | 9.83 |  | 7.56 |  | 16.39 |  | 4.17 |  | 47.9 |  | 58.2 |  | 63.9 |  | 60.3 |  |
| Practises SH | 5.75 | .095 | 10.78 | .718 | 8.00 | .954 | 17.75 | .299 | 3.63 | .882 | 50.5 | .970 | 62.1 | .151 | 65.5 | .717 | 61.9 | .470 |
| Does not practise SH | 9.55 |  | 10.19 |  | 8.10 |  | 15.45 |  | 3.75 |  | 50.3 |  | 53.9 |  | 62.6 |  | 58.1 |  |
| 2+ awakenings /night | 9.27 | .412 | 10.31 | .938 | 7.25 | .240 | 17.94 | .028 | 3.50 | .514 | 48.7 | .530 | 56.6 | .858 | 66.1 | .388 | 58.3 | .668 |
| 0-1 awakenings/night | 7.54 |  | 10.43 |  | 9.00 |  | 13.67 |  | 4.00 |  | 52.5 |  | 55.7 |  | 59.8 |  | 60.3 |  |
| Nappers* | 11.86 | .043 | 8.43 | .074<sup>d</sup> | 9.00 | .766 | 15.43 | .371 | 4.00 | .261 | 42.4 | .110 | 55.6 | .931 | 56.0 | .402 | 59.0 | .824 |
| Non-nappers | 6.86 |  | 10.64 |  | 8.43 |  | 17.57 |  | 3.07 |  | 54.1 |  | 56.1 |  | 63.7 |  | 57.7 |  |
| Uses alarm clock | 10.83 | .044 | 9.00 | .126 | 8.00 | .942 | 16.58 | .683 | 4.00 | .545<sup>d</sup> | 45.9 | .199 | 57.8 | .586<sup>d</sup> | 64.6 | .779 | 66.0 | .008 |
| Does not use alarm clock | 6.69 |  | 11.28 |  | 8.11 |  | 15.75 |  | 3.50 |  | 53.6 |  | 55.1 |  | 62.5 |  | 54.0 |  |
| Obese | 9.54 | .340 | 10.50 | .867 | 7.38 | .322 | 15.14 | .335 | 4.71 | .005 | 44.0 | .028 | 53.9 | .386 | 59.5 | .285 | 57.4 | .474 |
| Non-obese | 7.53 |  | 10.25 |  | 8.86 |  | 17.07 |  | 2.71 |  | 56.7 |  | 58.5 |  | 67.3 |  | 60.9 |  |
| Overweight + Obese | 8.79 | .446 | 10.24 | .704 | 8.52 | .171 | 16.33 | .582 | 3.92 | .183 | 49.6 | .571 | 56.9 | .541 | 62.8 | .709 | 59.6 | .653 |
| Normal BMI | 6.5 |  | 11.0 |  | 5.8 |  | 14.75 |  | 2.50 |  | 54.5 |  | 52.3 |  | 66.8 |  | 56.5 |  |
| Hypertensive | 7.93 | .589 | 10.25 | .863<sup>d</sup> | 7.38 | .322 | 15.8 | .744 | 4.67 | .004 | 48.8 | .587 | 50.8 | .373 | 58.4 | .198 | 59.6 | .855 |
| Normotensive | 9.08 |  | 10.50 |  | 8.86 |  | 16.46 |  | 2.62 |  | 52.1 |  | 53.7 |  | 67.7 |  | 58.71 |  |
| MM classes 3-4 | 9.44 | .522 | 11.33 | .393 | 7.78 | .802 | 18.22 | .141 | 5.33 | .001 | 41.8 | .042 | 54.2 | .605 | 63.9 | .926 | 59.0 | .967 |
| MM classes 1-2 | 8.00 |  | 9.95 |  | 8.19 |  | 15.11 |  | 2.95 |  | 54.4 |  | 57.2 |  | 63.2 |  | 59.21 |  |
| Abnormal WC<sup>‡</sup> | 8.76 | .625 | 10.50 | .767 | 8.27 | .649 | 16.24 | .822 | 4.05 | .022<sup>d</sup> | 46.9 | .038 | 56.1 | .914 | 64.7 | .547 | 58.2 | .513 |
| Normal WC | 7.57 |  | 10.00 |  | 7.50 |  | 15.71 |  | 2.17 |  | 60.7 |  | 56.7 |  | 53.6 |  | 61.9 |  |
| Abnormal NC<sup>◊</sup> | 9.08 | .611 | 10.69 | .702 | 8.15 | .919 | 15.67 | .639 | 4.85 | .005<sup>d</sup> | 44.9 | .082 | 58.4 | .428 | 68.5 | .171<sup>d</sup> | 59.5 | .878 |
| Normal NC | 8.00 |  | 10.12 |  | 8.00 |  | 16.62 |  | 2.73 |  | 55.1 |  | 54.3 |  | 58.9 |  | 58.8 |  |
| WHOQOL-Bref** |  |  |  |  |  |  |  |  |  |  |  |  |  |  |  |  |  |  |
| • D1 ≥ 60% | 8.67 | .843 | 10.12 | .743 | 8.12 | .753 | 16.68 | .092 | 3.80 | .514 | — | — | — | — | — | — | — | — |
| • D1 < 60% | 9.33 |  | 9.33 |  | 7.33 |  | 11.33 |  | 3.00 |  | — | — | — | — | — | — | — | — |
| • D2 ≥ 60% | 8.58 | .814 | 10.40 | .432 | 8.50 | .338 | 15.90 | .746 | 3.45 | .267 | — | — | — | — | — | — | — | — |
| • D2 < 60% | 9.13 |  | 9.3 |  | 6.88 |  | 16.63 |  | 4.38 |  | — | — | — | — | — | — | — | — |
| • D3 ≥ 60% | 7.92 | .484 | 10.00 | .967 | 9.58 | .074 | 17.25 | .323 | 3.58 | .766 | — | — | — | — | — | — | — | — |
| • D3 < 60% | 9.40 |  | 10.06 |  | 6.88 |  | 15.25 |  | 3.81 |  | — | — | — | — | — | — | — | — |
| • D4 ≥ 60% | 7.23 | .161 | 11.07 | .155 | 9.07 | .173 | 16.64 | .595 | 3.79 | .851 | — | — | — | — | — | — | — | — |
| • D4 < 60% | 10.14 |  | 9.00 |  | 7.00 |  | 15.57 |  | 3.64 |  | — | — | — | — | — | — | — | — |

**Table 3.** independence Chi-Square and exact Fisher tests for different groups or conditions, considering ESS and ISI as categorical attributes (people with EDS vs. with non-EDS and insomniacs vs. non-insomniacs, respectively, with cut-off values of 9 for ESS and 15 for ISI) * N = 21, ** N = 28, f – Fisher’s exact test was performed in this situation (otherwise, Chi squared test was performed)

| Condition/Group | ESS < 11 |  | ESS ≥ 11 |  | p | ISI < 15** |  | ISI ≥ 15** |  | p |
| --- | --- | --- | --- | --- | --- | --- | --- | --- | --- | --- |
|  | N |  | N | % |  | N |  | N | % |  |
| Snorer | 3 |  | 4 | 57,1 | .181<sup>f</sup> | 4 |  | 3 | 42.9 | .666 |
| Non-snorer | 17 |  | 6 | 26,1 |  | 9 |  | 14 | 60.9 |  |
| Insomnia | 15 |  | 4 | 21,1 | .108<sup>f</sup> | 7 |  | 12 | 63.2 | .454<sup>f</sup> |
| Non-Insomnia | 5 |  | 6 | 54,5 |  | 6 |  | 5 | 45.5 |  |
| Abnormal WC | 6 |  | 2 | 25.0 | .682<sup>f</sup> | 8 |  | 14 | 63.6 | .175<sup>f</sup> |
| Normal WC | 14 |  | 8 | 36.4 |  | 5 |  | 3 | 37.5 |  |
| Abnormal NC | 11 |  | 6 | 35.3 | 1.0<sup>f</sup> | 10 |  | 16 | 61.5 | .242<sup>f</sup> |
| Normal NC | 9 |  | 4 | 30.8 |  | 2 |  | 0 | 0 |  |
| Obese | 8 |  | 6 | 42.9 | .442<sup>f</sup> | 5 |  | 9 | 64,3 | .431 |
| Non-obese | 12 |  | 4 | 25.0 |  | 8 |  | 8 | 50.0 |  |
| Overweight + Obese | 15 |  | 10 | 40.0 | .140<sup>f</sup> | 10 |  | 15 | 60.0 | .628<sup>f</sup> |
| Normal BMI | 5 |  | 0 | 0.0 |  | 3 |  | 2 | 40.0 |  |
| MM grades 3-4 | 6 |  | 3 | 33.3 | 1.0<sup>f</sup> | 2 |  | 7 | 77,8 | .229<sup>f</sup> |
| MM grades 1-2 | 14 |  | 7 | 33.3 |  | 11 |  | 10 | 47.6 |  |
| 2+ awakenings /night | 12 |  | 5 | 29.4 | .705<sup>f</sup> | 4 |  | 12 | 75.0 | .121<sup>f</sup> |
| 0-1 awakenings/night | 8 |  | 5 | 38.5 |  | 7 |  | 5 | 41.7 |  |
| HADS-A ≥ 12 | 10 |  | 2 | 16.7 | .235<sup>f</sup> | 5 |  | 7 | 58.3 | .880 |
| HADS-A < 12 | 10 |  | 8 | 44.4 |  | 8 |  | 10 | 55.6 |  |
| HADS-D ≥ 12 | 4 |  | 3 | 42.9 | .657<sup>f</sup> | 2 |  | 5 | 71.4 | .427<sup>f</sup> |
| HADS-D < 12 | 16 |  | 7 | 30.4 |  | 11 |  | 12 | 52.2 |  |
| STOP-BANG ≥ 5 | 3 |  | 5 | 62.5 | .078<sup>f</sup> | 3 |  | 5 | 62.5 | 1.0<sup>f</sup> |
| STOP-BANG < 5 | 17 |  | 5 | 22.7 |  | 10 |  | 12 | 54.5 |  |
| Alcohol drinkers | 11 |  | 6 | 35.3 | .547 | 7 |  | 10 | 58.8 | .785 |
| Non-alcohol drinkers | 11 |  | 6 | 35.3 |  | 6 |  | 7 | 53.8 |  |
| Hypertensives | 11 |  | 5 | 31.3 | 1.0<sup>f</sup> | 7 |  | 9 | 56.3 | .961 |
| Normotensives | 9 |  | 5 | 35.7 |  | 6 |  | 8 | 57.1 |  |
| Diabetic/Prediabetic | 6 |  | 4 | 40.0 | .633<sup>f</sup> | 1 |  | 4 | 80.0 | .619<sup>f</sup> |
| Non-(Pre)Diabetic | 15 |  | 9 | 37.5 |  | 10 |  | 13 | 56.5 |  |
| Uses alarm clock | 5 |  | 7 | 58.3 | .045<sup>f</sup> | 3 |  | 9 | 75.0 | .098 |
| Doesn't use alarm clock | 15 |  | 3 | 16.7 |  | 10 |  | 8 | 44.4 |  |
| Uses smartwatch | 3 |  | 2 | 40.0 | 1.0<sup>f</sup> | 0 |  | 4 | 100 | .132<sup>f</sup> |
| Does not use smartwatch | 17 |  | 8 | 32.0 |  | 11 |  | 13 | 21.4 |  |
| Exercises | 9 |  | 3 | 25.0 | .694<sup>f</sup> | 5 |  | 7 | 58.3 | .880 |
| Sedentary | 11 |  | 7 | 38.9 |  | 8 |  | 10 | 55.6 |  |
| Practises SH | 6 |  | 3 | 33.3 | .704<sup>f</sup> | 2 |  | 7 | 77.8 | .229<sup>f</sup> |
| Doesn't practise SH | 12 |  | 9 | 42.9 |  | 11 |  | 10 | 47.6 |  |
| Nappers* | 3 |  | 4 | 57.1 | .102<sup>f</sup> | 2 |  | 5 | 71.4 | .642<sup>f</sup> |
| Non-nappers* | 11 |  | 3 | 21.4 |  | 7 |  | 7 | 50.0 |  |
| WHOQOL BREF** |  |  |  |  |  |  |  |  |  |  |
| D1 ≥ 60% | 4 |  | 7 | 63.6 | .020<sup>f</sup> | 2 |  | 1 | 33.3 | .543<sup>f</sup> |
| D1 < 60% | 14 |  | 3 | 17.6 |  | 9 |  | 16 | 64.0 |  |
| D2 ≥ 60% | 4 |  | 2 | 33.3 | 1.0<sup>f</sup> | 4 |  | 4 | 50.0 | .671<sup>f</sup> |
| D2 < 60% | 14 |  | 8 | 36.4 |  | 7 |  | 13 | 65.0 |  |
| D3 ≥ 60% | 11 |  | 5 | 31.3 | .687<sup>f</sup> | 7 |  | 9 | 56.3 | .705<sup>f</sup> |
| D3 < 60% | 6 |  | 5 | 45.5 |  | 4 |  | 8 | 66.7 |  |
| D4 ≥ 60% | 7 |  | 7 | 50.0 | .115 | 6 |  | 8 | 57.1 | .699 |
| D4 < 60% | 11 |  | 3 | 21.4 |  | 5 |  | 9 | 64.3 |  |

Some conditions, although inquired/inspected by the researchers, could not be further scrutinized due to reduced sample size - we did not run statistics when a group was smaller than 5 subjects - and, therefore, with low statistical power, such as smokers (only 3) or jaw misalignments (only 2 retrognathic and 2 prognathic subjects). We also could not divide further groups into subgroups due to the same reason, as for instance separation, between those who exercised, according to preferred time of practice (diurnal vs. nocturnal) or according to the modality of the exercise (aerobic vs. anaerobic) etc.

## DISCUSSION

There were relevant associations on **Table 2**, regarding ESS, ISI, STOP-BANG and WHOQOL-BREF, but not concerning HADS.

With respect to ESS, women were significantly associated with lower scores, and daytime nappers and people who woke with the aid of an alarm clock were associated with higher scores.

On the other hand, ISI scores were significantly higher only in those who woke 2 or more times per night on average, in contrast to those who woke only once or did not wake at all.

As to STOP-BANG, sedentary, obese (but not overweight), those with unfavourable respiratory airway (MM of classes 3 and 4), and hypertensive patients had significant higher scores than correlates. People with broader neck circumferences and obese also had a positive correlation, but are already part of the scoring in this questionnaire - although, in terms of body mass, only BMI ≥ 35, (i.e., Classes II and III obesity), counts as one point when scoring STOP-BANG, and in our series, we included people with BMI ≥ 30 to also embrace Class I obese, so that behaviour was expected.

WHOQOL-BREF had statistically significant associations only for the first and fourth domains, which represent Physical and Environmental wellbeing, respectively. Non-obese, people with more favourable airway (MM Classes I and II) and people with normal WC scored significantly higher than their counterparts on D1, while men and people who relied on an alarm clock to wake up performed quite better than women and those who did not use alarm clock, in that order.

Both Tables 2 and 3 frequently show greater scores (in Table 2) or greater tendency to be more frequent classified in daytime sleepers or insomniacs (Table 3) in groups that were expected to perform worse, but in many times - especially in Table 3, they failed to reach statistical significance - examples: In Table 2, Women’s mean HADS-D score was 11.3, while men’s was 8.9, 57% EDS was found in snorers vs. 26% in non-snorers, or insomnia symptoms in 75% of those who awoke 2 or more times at night vs. 42% in those who did not - only the association of alarm clock and higher scores of D1 were associated to a level of p < .05.

We believe that the small sample contributed negatively towards statistical significance for the bulk of cases. Nonetheless, as said, in most part the results are in accordance to what was expected both by experience of the authors and by vast, previous literature results.

## CONCLUSION

This is a transversal survey that intends to bring to light some knowledge of the profile of sleep complaints in our local population. In accordance to our literature review, the main complaints were insomnia and snoring/witnessed apnoea.

Sleep complaints were associated with worsening of the quality of life in our sample, especially on the physical and environmental domains, according to the WHOQoL-BREF. Our sample reflects a local population with a considerable rate of sleep issues, and serves as an alert for the importance of public health policies targeting conscientization of the population and health workers for caring about the sleep health.

## Data Availability

All data produced in the present work are contained in the manuscript

